# Autism in gifted youth is associated with low processing speed and high verbal ability

**DOI:** 10.1101/2021.11.02.21265802

**Authors:** Jacob J. Michaelson, Alissa Doobay, Lucas Casten, Katherine Schabilion, Megan Foley-Nicpon, Thomas Nickl-Jockschat, Ted Abel, Susan Assouline

**Affiliations:** Department of Psychiatry, University of Iowa, Iowa City, IA, USA; Iowa Neuroscience Institute, Carver College of Medicine, University of Iowa, Iowa City, IA, USA; Belin-Blank Center for Gifted Education and Talented Development, College of Education, University of Iowa, Iowa City, IA; Department of Psychological and Quantitative Foundations, University of Iowa, Iowa City, IA, USA; Department of Neuroscience and Pharmacology, Carver College of Medicine, University of Iowa, Iowa City, IA, USA

## Abstract

**Background:** High cognitive ability is an almost universally positive prognostic indicator in the context of neurodevelopmental, neuropsychiatric, and neurodegenerative conditions. However, “twice-exceptional” individuals, those who demonstrate exceptionally high cognitive ability (gifted) and exhibit profound behavioral and mental health challenges, are a striking exception to this rule.

**Methods:** We digitized the clinical records of N=1,074 clients from a US-based specialty clinic serving gifted students. This included a broad array of diagnostic, cognitive, achievement, and behavioral data, including self, teacher, and parent reported items. We conducted both hypothesis-driven and unsupervised learning analyses to 1) identify characteristics whose association with full-scale IQ (FSIQ) was dependent on autism diagnosis and 2) identify cognitive archetypes associated with autism diagnosis and related behaviors. We tested the generalization of our findings using data from the SPARK (N=17,634) and ABCD studies (N=10,602).

**Results:** Autistic individuals with IQ >= 120 were nearly 15 times more likely to enter adulthood undiagnosed compared to lower-IQ (IQ < 70) counterparts. Self-reported sense of inadequacy was most strongly associated with increasing FSIQ specifically among autistic clients (beta=0.3, 95% CI:[0.15,0.45], p=7.1×10^−5^). Similarly, self, parent, and teacher reports of anxiety increased with FSIQ (all p<0.05) in autistic individuals, in striking opposition to the ameliorating effect of FSIQ seen in non-autistic individuals. We uncovered a pattern of decreased processing speed (PS) coupled with very high verbal comprehension (VC), a PS/VC discrepancy, that was associated with autism, inattention, and internalizing problems. Similar cognitive-behavioral links were also observed in the ABCD study. Finally, we found a significant association between the PS/VC discrepancy and polygenic risk for autism in the ABCD sample (t=2.9, p=0.004).

**Conclusions:** Our results suggest that autistic individuals with exceptional ability are underserved and suffer disproportionately from high anxiety and low self-worth. In addition, elevated IQ with a significant PS/VC discrepancy appears to be a clinically and genetically meaningful biotype linked to autism.

## INTRODUCTION

The debate about whether giftedness (i.e., extremely high intelligence) is a protective or risk factor for neuropsychiatric disorders is nearly as old as the field of psychiatry itself [1–3]. While some studies have reported a potential association for high ability with neurodevelopmental and psychiatric disorders [4, 5], and, in particular, with genetic risk for those disorders [6–9], the prevalent notion in the field is that higher IQ is a protective factor [3, 10]. Both possibilities – extreme cognitive ability either being a protective or a predisposing factor – could hint at distinct underlying neurobiological mechanisms. However, this apparent paradox highlights how the shared neurobiology underlying intelligence and psychiatric risk is still poorly understood.

Although measurements of intelligence have evolved since their first documented uses, one consistent theme of these assessments is the idea that general intelligence is comprised of distinct cognitive skill domains [11]. These domains typically include verbal comprehension (VC), perceptual reasoning (PR), processing speed (PS), working memory (WM), and a composite score called a Full-Scale Intelligence Quotient (FSIQ). Studies of these scores have found that, overall, increased FSIQ is related to better outcomes and lower rates of psychiatric conditions and mental illness [12], but deficits in specific domains are related to an increased rate of psychiatric diagnoses [13].

Two specific intelligence domains of special interest to neurodevelopmental and psychiatric disorders are PS and VC. PS is typically defined as how efficiently the brain can take sensory (e.g., visual) input, accurately decode that information, and produce a response. Measurement typically includes a proctored test involving a “key” set of patterns, with the subject identifying as many matches as possible in a short time period [14]. VC is commonly defined as the ability to understand concepts and effectively communicate [14]. Measurement of VC typically involves assessing vocabulary, ability to abstract similar words, and ability to understand complex expressions [14]. Studies of PS in autism have consistently found deficits across cohorts not seen in other neurodevelopmental conditions [13, 15]. Conversely, similar studies have sometimes found relative strengths in VC scores across neurodevelopmental and psychiatric disorders, including autism and bipolar disorder [15–19]. However, the frequent co-morbidity of intellectual disability in some studies may contribute to inconsistent observation of a specific VC strength.

At the most extreme end of the cognitive discrepancy spectrum are “twice-exceptional” (2e) individuals, defined as having any IQ domain score greater than 120 (approximately the 91^st^ percentile) and a co-morbid neuropsychiatric diagnosis, like autism [20, 21]. Thus, 2e individuals may be an example of a group where high IQ, particularly when it is unevenly distributed across domains of intelligence, may indicate a neurodevelopmental liability. Studying the relative strengths and weaknesses of 2e individuals with autism could provide insights into how deficits in one specific intelligence domain may reflect disrupted neurobiology and identify pathways key to cognition and psychiatric risk.

Further evidence the neurobiology of intelligence and autism is intertwined comes from genetic studies. Twin studies of IQ have found it is one of the most heritable brain related traits, with a peak heritability estimate of ~0.8 in adulthood [22]. Similarly high heritability estimates have been seen in autism, which is currently estimated at 0.8 [23]. Genome Wide Association Studies (GWAS) for proxies of intelligence, like years of education, have identified over 1,200 associated genetic loci [24]. Examination of the genes implicated by these associated loci showed significant functional enrichment for processes known to be disrupted in autism, particularly in pathways related to dendrite morphology and neuronal migration [24]. Analyses of GWAS associations in intelligence and autism have found the two traits are significantly genetically correlated with each other [7]. Better characterization of the specific domains of intelligence impacted in autism could provide meaningful insights into the neurobiology underlying both autism and intelligence.

Here, we present what we understand to be the largest study yet of the intersection of giftedness (i.e., extremely high IQ) and autism, and we reveal a specific cognitive and behavioral profile that is over-represented among these twice-exceptional individuals. Furthermore, we find that elements of this cognitive-behavioral profile are associated with polygenic risk for autism in a general population sample. Specifically, this study aims to answer the following questions: What behaviors, if any, become more problematic for autistic individuals with increasing IQ? What effect does IQ have on age at diagnosis, a factor critical for granting affected individuals access to specialized care? Does an axis of discrepancy between PS and VC discriminate autism from typically developing individuals? What are the behavioral correlates of the PS/VC discrepancy? Does the PS/VC axis predict similar behavioral issues in non-autistic individuals? Finally, what are the genetic correlates of this cognitive ability discrepancy? Answering these questions necessitates the study of twice-exceptional individuals with autism, who are likely to show more extreme cognitive discrepancies in PS and VC than autistic cohorts sampled without attention to IQ.

## METHODS AND MATERIALS

### Cohort 1: gifted clinical sample

Over a ten-year period (2009-2019), N=1,074 children and adolescents ages six to 18 were evaluated at a university-based clinic that specializes in the assessment and counseling of gifted and twice-exceptional students. Some of the students were evaluated more than one time (1,254 total visits). Although students in the sample had Full Scale IQs as high as 158, a few of the clients evaluated at the clinic had very low IQs (e.g., 55). The average IQ for the cohort was 116.9 (SD of 14.5), the median was 117. Some individuals in the cohort were diagnosed with ASD, whereas others received a different diagnosis such as ADHD or SLD, and many did not have a diagnosis. Thus, from the perspective of cognitive ability, this cohort provided a broader IQ distribution compared to previously cited twice-exceptional research[17–19] in which the samples only included autistic individuals with very high IQ (twice-exceptional), i.e., IQ ≥ 120. Consequently, the current study offers a more inclusive IQ and diagnostic sample: autistic individuals within the average range, or below, individuals with very high IQ and not diagnosed with ASD (i.e., gifted only), and non-autistic individuals with IQs in the average range. Demographic information concerning these participants is presented in Table 1.

**Table 1:** Demographics of the Belin-Blank Center gifted youth sample (N=1,074).

| Dx group | male (%) | Ethnicity: white (%) | Ethnicity: 2 or more (%) | Asian of Pacific Islander (%) | Hispanic or Latin(x) (%) | African American or Black (%) | American Indian (%) | Age in years (s.d.) |
| --- | --- | --- | --- | --- | --- | --- | --- | --- |
| all | 68.44 | 89.62 | 3.67 | 3.67 | 2.41 | 0.52 | 0.1 | 10.16 (3.7) |
| ASD | 83.33 | 88.79 | 3.45 | 4.31 | 3.45 | 0 | 0 | 10.8 (3.91) |
| ALD | 70.52 | 89.17 | 7.64 | 1.91 | 1.27 | 0 | 0 | 10.71 (2.89) |
| ADHD | 74.65 | 91.49 | 2.74 | 1.82 | 3.04 | 0.61 | 0.3 | 10.75 (3.5) |
| none | 63.07 | 88.74 | 1.99 | 6.62 | 1.66 | 0.99 | 0 | 9.25 (3.6) |
| other | 62.8 | 90.26 | 4.62 | 2.56 | 2.56 | 0 | 0 | 10.47 (3.98) |

### Cohort 2: SPARK

SPARK[25] is an ongoing, US-based genetic study of autism of nearly 300,000 individuals. In the current study, we used SPARK data primarily to understand the relationship between age at diagnosis (provided during enrollment) and IQ. In SPARK, two indices of IQ are available for a subset of participants: 1) parent-reported IQ in bins of approximately 10 points (N=17,634) and 2) IQ scores as contained in the electronic health record of participants and uploaded into a central SPARK database by the clinical recruitment centers (N=3,339). For both of these IQ indices, we obtained data from the SPARK phenotype release V7. SPARK is approved by the Western IRB (IRB 20151664). All participants provided informed consent.

### Cohort 3: ABCD

The Adolescent Brain Cognitive Development (ABCD) study [26] is a general population sample that does not exclude on the basis of neuropsychiatric diagnoses. The study includes longitudinal data collection across behavioral and neuroimaging modalities and also includes genetic data. For cognitive-behavioral analyses, release 2 data were used (N=10,601). For genetic analyses, the cohort was further filtered to participants in which genetic data were available that passed our quality control and clustered in the majority European cluster based on SNPs (genetic clustering detailed below). After this filtering, 6,559 ABCD participants remained. The ABCD cohort was genotyped on the Affymetrix NIDA SmokeScreen Array and was processed through standard QC steps before release, including removing SNPs with low call rate and individuals with potential contamination problems or high missing data. The SNP QC process was based on the recommendations by [27] using PLINK [28] and R [29] with the same parameters used for the SPARK genotyping QC. In total, 399,016 SNPs and 9,324 individuals passed QC. After this QC, the remaining cohort was merged and clustered with the 1,000 Genomes Phase 3. Clustering was based on the first 10 components from multi-dimensional scaling of the combined kinship matrix of the cohort and 1,000 Genomes. This combined cohort was clustered into 5 groups, representing the 5 distinct super-populations. For genotype imputation, ambiguous SNPs were also removed, leaving 372,694 SNPs. These remaining individuals and SNPs were imputed to the 1,000 Genomes Phase 3 ALL reference panel using the Genipe pipeline [30]. 6,659 individuals clustered in the European cluster. Genipe performed LD calculation and pruning with PLINK [28], genotype phasing with SHAPEIT [31], and genotype imputation by IMPUTE2 [32] using default parameters.

### Psychoeducational and diagnostic instruments

Sattler [33] offers a thorough description of the Wechsler Intelligence Scale for Children (5th Edition; WISC-V). Multiple subtests comprise the WISC-V, including ten primary subtests: Block Design, Similarities, Matrix Reasoning, Digit Span, Coding, Vocabulary, Figure Weights, Visual Puzzles, Picture Span, and Symbol Search. From the ten primary subtests, the first seven are used to obtain the Full Scale IQ (FSIQ). In addition to the ten primary subtests, there are six secondary subtests as well as five complementary subtests. Scores from the Similarities and Vocabulary subtests comprise the Verbal Comprehension Index; Block Design and Visual Puzzles subtest scores comprise the Visual Spatial Index; Matrix Reasoning and Figure Weights subtest scores comprise the Fluid Reasoning Index; Digit Span and Picture Span comprise the Working Memory Index, and Coding and Symbol Search comprise the Processing Speed Index. The primary index scores as well as the FSIQ, are reported as standard scores with a mean of 100 and a standard deviation of 15. Standard scores for each of the scales have a mean score of 10 and a standard deviation of 3.

The WISC-V [34] is a widely used measure of intellectual ability with good psychometric properties (see the WISC-V Technical and Interpretive Manual; [35]). Confirmatory factor analysis shows that the WISC-V measures five interrelated but distinct general abilities; thus, empirical data match the hypothesized structure of the test, which is rooted in contemporary intelligence theory [34]. Criterion validity studies demonstrate that the WISC-V can validly be interpreted as a measure of intelligence in children based on intercorrelations with the Kaufman Assessment Battery for Children (KABC-II; the WISC-V FSIQ and KABC-II Fluid Crystallized Index and Mental Processing Index were 0.77 to 0.81, respectively), the Kaufman Test of Educational Achievement (KTEA-3; most correlations were found to be within the Moderate range), the Wechsler Individual Achievement Test (WIAT-III; moderate correlations ranging from 0.40 to 0.73 were found between the WISC-5 FSIQ and WIAT-3 Composite scores), and the Wechsler Abbreviated Scale of Intelligence (WASI-II; correlation coefficients are moderately high, ranging from 0.57 to 0.87). Test-retest reliability of the WISC-V is high, ranging from 0.88 (Processing Speed) to 0.96 (FSIQ and GAI). WISC-5 internal consistency estimates range from 0.81 to 0.94; for the FSIQ, internal consistency across the 11 age groups ranged from 0.96 to 0.97.

The Behavior Assessment System for Children, 3rd Edition (BASC-3; [36]) is described as “a multidimensional system used to evaluate the behavior and self-perceptions of children and young adults ages 2 through 25 years” (p.1). There are multiple components, including a self-report (SRP), a teacher-report (TRS), and a parent-report (PRS). Each of the three forms is further divided into Pre-school (age 2-5), Child (ages 6 through 11 years), Adolescent (ages 12 through 18 years), and College (18-25) forms that contain similar subscales to facilitate comparisons between forms; for 6-to 7-year-old students, the self-report form is administered as an interview (SRP-I). Scores are reported as T-scores with a mean of 50 and standard deviation of 10. There are two types of scales: clinical scales (measuring behavioral and emotional concerns) and adaptive scales (measuring strengths or protective factors). Among the clinical scales, scores of 60 through 69 are considered At-Risk and scores of 70 and above are considered Clinically Significant. Among the adaptive scales, scores of 31 through 40 are considered At-Risk, whereas scores of 30 and below are considered Clinically Significant.

There is extensive research supporting the validity and reliability of the BASC-3 in clinical settings [36]. Reliability measures across the TRS, PRS, and SRP suggest good internal consistency for both scale and composite scores. Median internal consistency for the BASC-3 TRS yielded alpha coefficients between .87 and .91 for the clinical and adaptive scales, and between .95 and .97 for the composite scales [36]. Median internal consistency for the BASC-3 PRS yielded alpha coefficients between 0.83 and 0.89 for the clinical and adaptive scales, and between 0.93 and 0.97 for the composite scales [36]. For the SRP, alpha coefficients were between 0.82 and 0.86 for the clinical and adaptive scales, and between 0.93 and 0.95 for the composite scales. Test-retest reliability was also high across the TRS, PRS, and SRP forms. Test-retest reliability for the TRS ranged from 0.87 to 0.91 for the clinical and adaptive scales, and from 0.95 to 0.97 for the composite scales. The PRS had test-retest reliabilities ranging from 0.83-0.89 for the clinical and adaptive scales, and between 0.93 and 0.97 for the composite scales. For the SRP, the clinical and adaptive scales had test-retest coefficients between 0.82 and 0.86, as well as composite scale coefficients between 0.93 and 0.95. The validity of the BASC-3 is substantially supported by the results of factor analyses consistent with scale composition, strong correlations with other instruments that assess emotional and behavioral symptoms in children (e.g., the Minnesota Multiphasic Personality Inventory and the Achenbach System of Empirically Based Assessment), and consistency between the results of this measure and the clinical diagnoses of the child being assessed. Additionally, there is good consistency between the BASC-3 and previous versions of the BASC, which have been used in hundreds of research studies in various settings, countries, languages, and clinical populations [36].

All BASC-3 clinical and adaptive skills were included in the present analysis. The BASC-3 scales of interest in this study included the TRS, PRS, and SRP Anxiety scale, the TRS School Problems scale, the PRS Functional Communication and Attention Problems Scales, and the SRP Sense of Inadequacy Scale and Hyperactivity scales.

### Cognitive ability and age at diagnosis

Data were pulled from the SPARK version 7 phenotype release (N=17,634 in this sample). Parents reported the approximate full-scale IQ (in bins spanning approximately ten points each) of their children as obtained through clinical testing. Age at professional diagnosis of autism was also reported by parents. We grouped these SPARK participants into three groups according to the parent-reported IQ: high (IQ ≥ 120), average (IQ 70-119) and low (IQ < 70). See Figure 2. Using age of six years as a threshold for diagnosis (corresponding with typical entry into the public school system), we compared diagnosis and non-diagnosis counts in the low and high IQ groups using a 2×2 table and Fisher’s exact test. Similarly, low or high IQ group membership was stratified by a binary “diagnosed by age 18” variable, indicating whether the individual would have been diagnosed by a child/adolescent or adult clinic. Odds ratios, their 95% confidence intervals, and p-values were recorded to assess significance of these tests of association.

**Figure 1.**
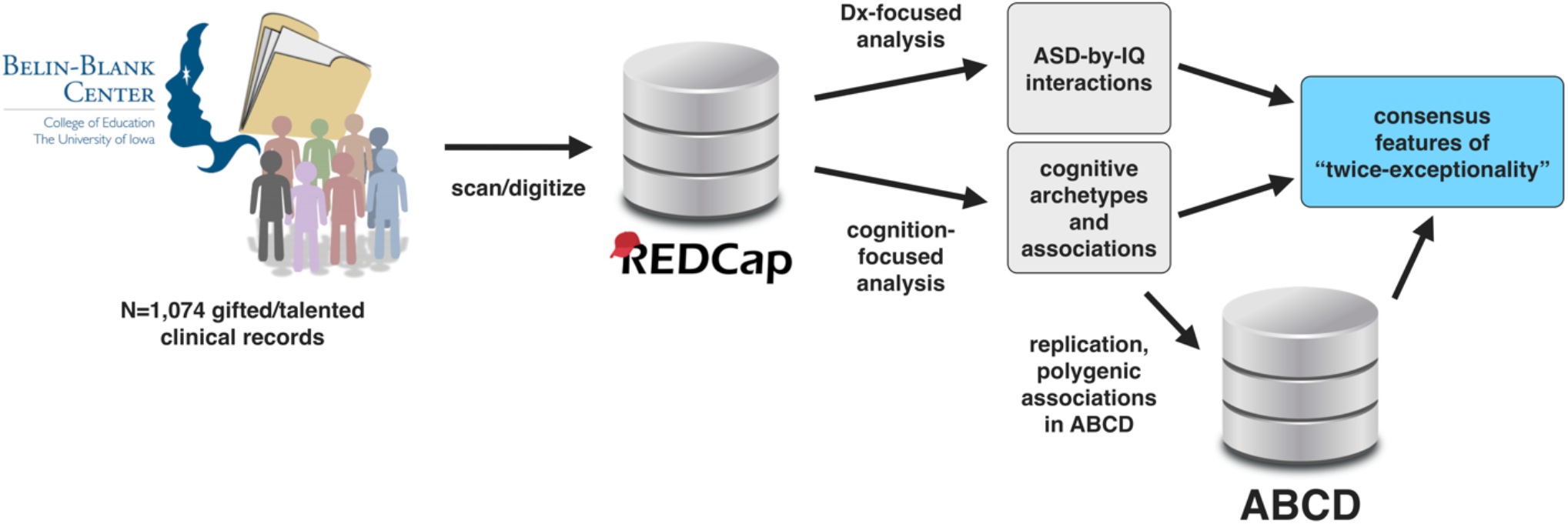
Overview of study design. Our discovery sample consisted of N=1,074 clients seen at the Belin-Blank Center (B-BC), a specialty clinic at the University of Iowa that serves gifted individuals. We digitized and entered the psychoeducational data from this sample into a REDCap database, which we then used to conduct hypothesis-driven and unsupervised learning analyses with the aim of gaining insight specifically into the cognitive, behavioral, and daily living challenges of “twice-exceptional” (2e) individuals: those with exceptional cognitive ability and having a diagnosis relevant to mental health. For the purposes of this study, we focus on autism, though twice-exceptionality extends beyond autism. Finally, to test the robustness and generalization of our findings, we used data from the ABCD study (N=10,601). We also used the genetic data in ABCD to compute polygenic risk scores for autism, which were then tested for association with traits of interest.

**Figure 2.**
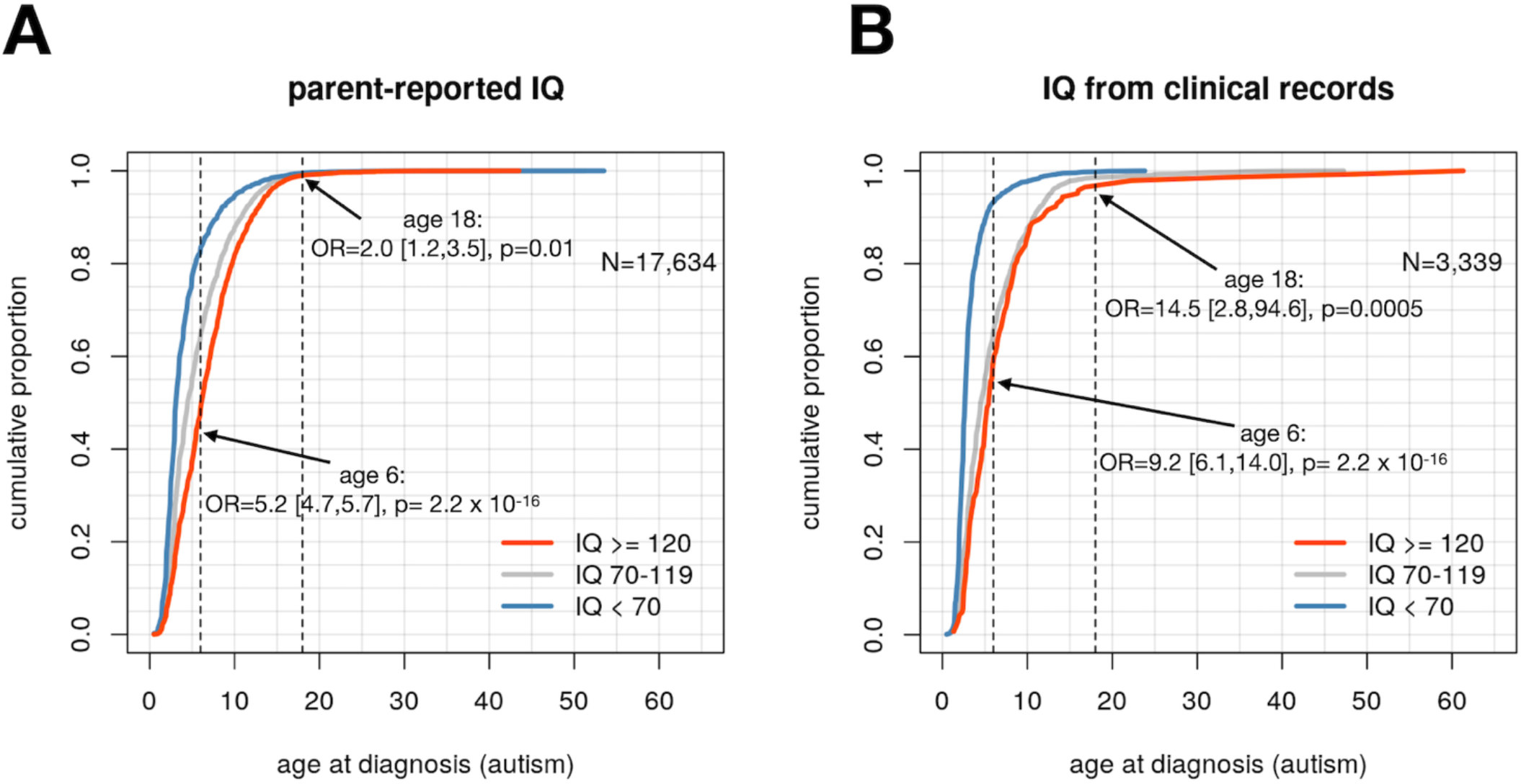
High IQ delays autism diagnosis. In a sample from the SPARK autism study, N=17,634 autistic individuals had a parent-reported IQ and age at autism diagnosis (A). To illustrate the relationship between IQ and age at diagnosis, we grouped these individuals into broad groups of low (< 70), average (70-119) and high (>= 120) IQ. Individuals in the high IQ group showed significantly delayed diagnosis compared to their low IQ counterparts, with high IQ individuals entering adulthood without a diagnosis at twice the rate of low IQ individuals, and entering school age (i.e., age 6 years) without a diagnosis at 5.2 times the rate of low IQ individuals. To address the sampling and reporting biases in this larger sample, we also examined the same comparisons in a smaller sample (N=3,339) with full scale IQ scores pulled from clinical records available to SPARK recruitment sites (B). These results follow the same overall trend as in A, but with much stronger effects: high IQ individuals enter adulthood without an autism diagnosis at 14.5 times the rate of their low IQ counterparts, and they enter school age (i.e., age 6 years) without a diagnosis at more than nine times the rate of their lower-IQ counterparts. Together, these results suggest significant delayed diagnosis and more importantly, crucial years of development spent without services and support for their disability.

**Figure 2.**
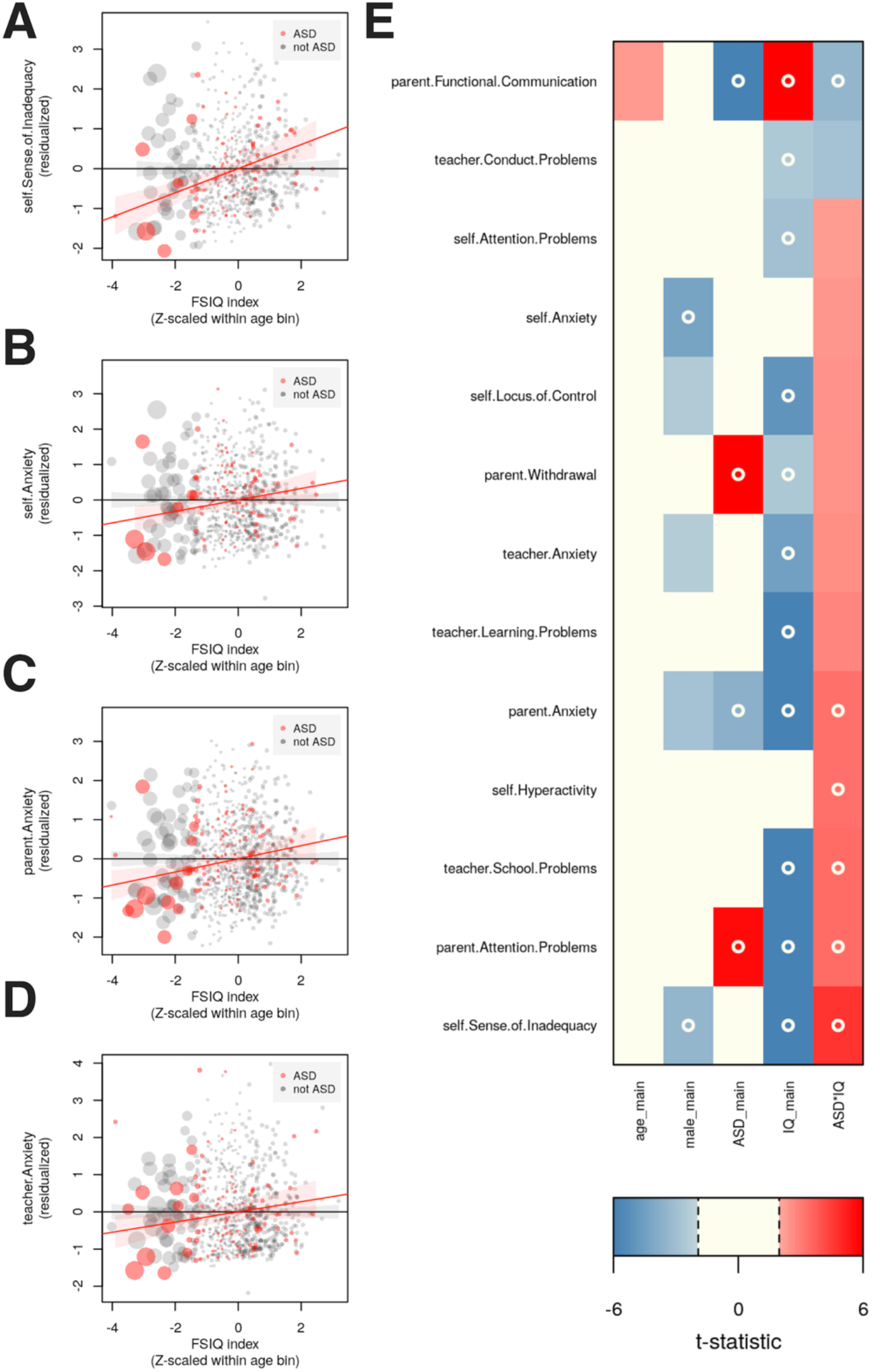
Interactions between IQ and ASD diagnosis are associated with specific anxiety and attention-related problems. Using psychoeducational data from clinical records, we tested scores from the BASC for association with IQ, ASD diagnosis, and their interaction (with sex and age as additional covariates). Most strikingly, the interaction between ASD and IQ leads to an increased sense of inadequacy (**A**) as IQ increases in those diagnosed with ASD (red points). Similarly, increasing IQ in autistic individuals is associated with increased anxiety according to self (**B**), parent (**C**), and teacher report (**D**). The remaining tests of association are summarized in (**E**), where t-statistics are shown, with FDR<0.05 indicated by a circle.

### Statistical analysis

Both BASC and IQ scores were standardized (mean 0 and SD 1) within each decile of age. For each measure, Analysis of Variance (ANOVA; anova() and lm() functions in R version 3.3) was utilized to test the significance of main effects of ASD diagnosis and full-scale IQ, as well as their interaction. In the linear model underlying the ANOVA, individuals were re-weighted such that for individuals with FSIQ < 120, their contribution to the least-squares fit was proportional to the pointwise estimate (based on their FSIQ) of density in a Gaussian distribution with mean 100 and SD 15. Collectively, individuals with FSIQ >= 120 were each weighted the same, and the sum of these weights equaled the sum of the weights for individuals with FSIQ < 120. This reweighting has the effect of emphasizing individuals whose FSIQ is close to the mean of 100, thus making the resulting comparisons with individuals with FSIQ > 120 more in line with what would be expected when comparing with a typical population sample. Although this approach had the effect of boosting power and resulted in a greater number of significant associations, in general the direction of association was congruent with an unweighted approach. The individual-level weights, together with the age-standardized IQ and BASC scores used in these analyses are included in Supplemental Table 1. For all ANOVA tests, adjustment for multiple testing was accomplished using the Benjamini-Hochberg procedure [37] and is reported as false discovery rate (FDR) where applicable.

### Unsupervised learning

We employed archetypal analysis [38] as implemented in the archetypes package for R. Input features were the Z-scaled index scores and subtest scores of the WISC-IV and WISC-V. These were imputed with the median value where missing (12% missingness). To minimize the impact of outliers, the archetypal analysis procedure was trained on 5,000 random mixtures of the original data, where the original data matrix was multiplied by a random mixing matrix following a Poisson distribution with rate=1. The values of this random mixing matrix were normalized such that the contribution from each of the 1,074 individuals summed to 1 for each mixture. The archetypal coefficients for the observed data were then inferred using the predict() function of the archetypes R package. The optimal number of archetypes was inferred using the elbow plot heuristic, which suggested diminishing returns after k=5 archetypes. Individuals were assigned to archetypes using the following heuristic: briefly, an individual is assigned to an archetype if the coefficient for that archetype is the maximal coefficient for that individual and it is more than twice the next-highest archetypal coefficient. Those individuals who did not meet this criterion were grouped together in a “mixed” group. Subsequent analyses used archetype coefficients as explanatory variables in linear models testing association with achievement and behavioral indices available in the database. To test association of the PS/VC discrepancy with BASC behavioral measures, we encoded a predictor variable as follows: 1 if the individual was assigned to A4, 3 if the individual was assigned to A5, and 2 for all others. This variable represents the ordinal progression from PS+/VC-(A4) to PS-/VC+ (A5). The models used for significance testing were therefore BASC ~ sex + FSIQ + PS_VC, where BASC is the measure in question from the BASC, FSIQ is the full-scale IQ, and PS_VC is the ordinal variable described above.

### Replication analysis

To determine whether key behaviors were robustly associated with a PS/VC discrepancy in an independent sample, we utilized data from the large ABCD study, a US-based general population study of adolescents with extensive behavioral, genetic, and neuroimaging data [39]. Specifically, we constructed a PS/VC “discrepancy score” by multiplying participants’ NIH toolbox scores for the pattern task (processing speed), the picture vocabulary task (verbal), and the reading task (verbal), by −1, 0.5, and 0.5, respectively, and then summing. These values were then Z-scaled to mean of 0 and standard deviation of 1. Individuals with large positive values of this score would have a large PS/VC-type discrepancy similar to that observed in A5 in the B-BC sample (Fig. 3A). Likewise, ABCD participants with an extreme negative score would have a similar discrepancy to individuals belonging to the A4 archetype in the B-BC sample (Fig. 3A). We note that there would have been a number of ways to approach this analysis (e.g., factor analysis, canonical correlation analysis, etc.). We investigated these alternative approaches and found that they yielded similar findings to those described below. However, given that the PS/VC discrepancy score based on the simple difference in task scores is more straightforward and directly interpretable, we chose that in the end. To identify behavioral correlates of the PS/VC discrepancy in ABCD, we tested associations between the ABCD PS/VC discrepancy score and subscales of the Child Behavior Checklist (CBCL)[40]. In total, twenty subscales derived from CBCL items are available in the ABCD data release (see Figure 4C). We tested linear models of the form y ~ sex + total_comp + PS_VC, where y is the CBCL subscale score under investigation, total_comp is the NIH Toolbox total composite score, and PS_VC is the PS/VC discrepancy score described above.

**Figure 3.**
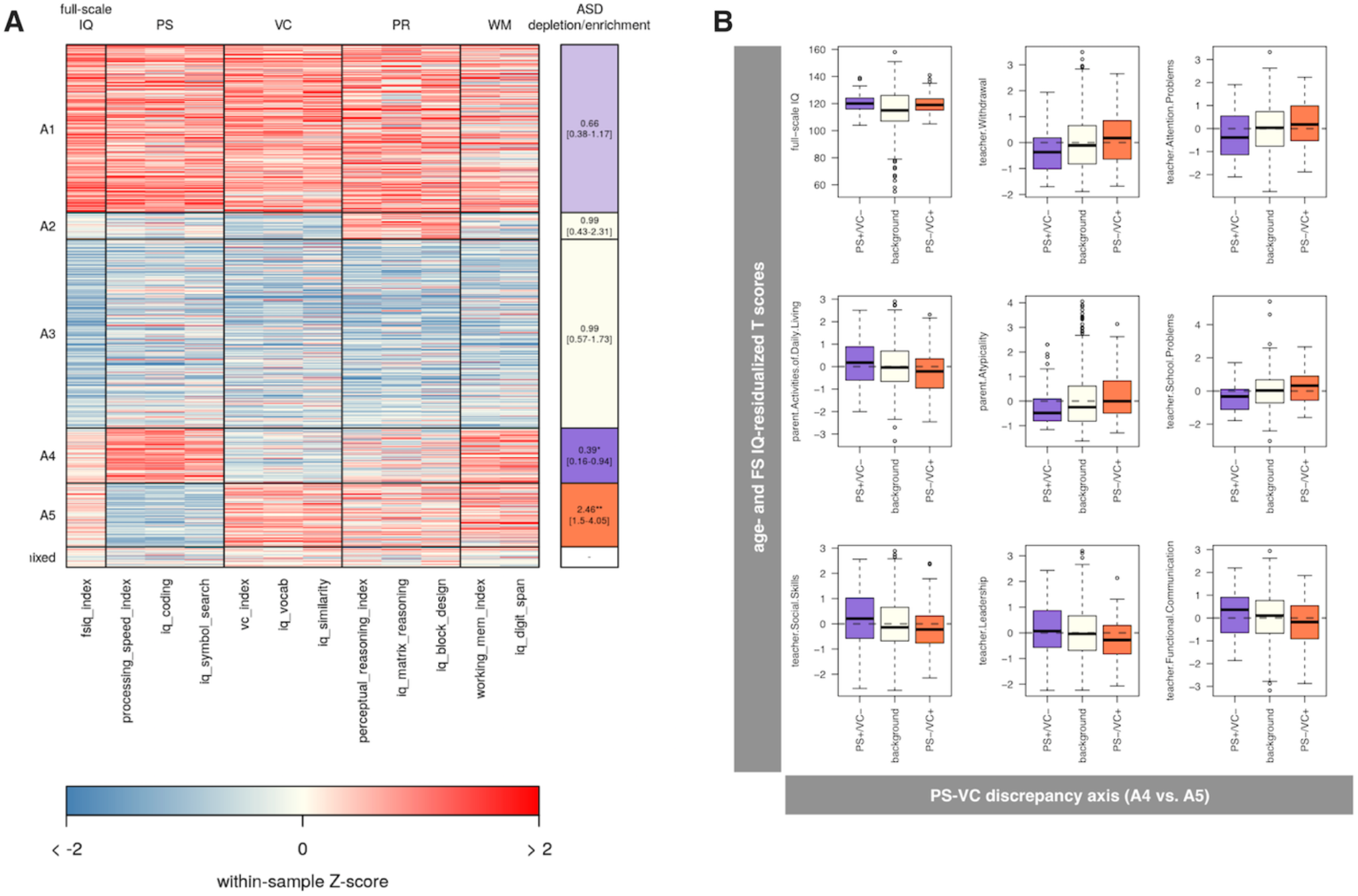
Unsupervised learning uncovers a discrepancy between processing speed (PS) and verbal comprehension (VC) that is significantly associated with autism diagnosis. We performed archetypal analysis, which yielded five distinct cognitive patterns or archetypes that best typify the patterns seen in the B-BC sample (**A**). In particular, archetype 4 (A4) and archetype 5 (A5) are nearly mirror-images of each other, with A5 showing a PS deficit and VC strength (PS-/VC+), and A4 showing a PS strength and VC deficit (PS+/VC-). A5 showed a significant over-representation of autistic individuals (OR=2.46, 95% CI: [1.5,4.1]), while A4 was significantly depleted (OR=0.39, 95% CI: [0.16,0.94]). Taken together, this suggests an axis of risk and resilience, defined by a PS/VC discrepancy. We tested subscales from the BASC for association with the PS/VC discrepancy (**B**) to identify specific problem areas linked to this cognitive pattern, highlighting attention and internalizing problems similar to those identified in the diagnosis-driven analysis (see **Figure 2**).

**Figure 4.**
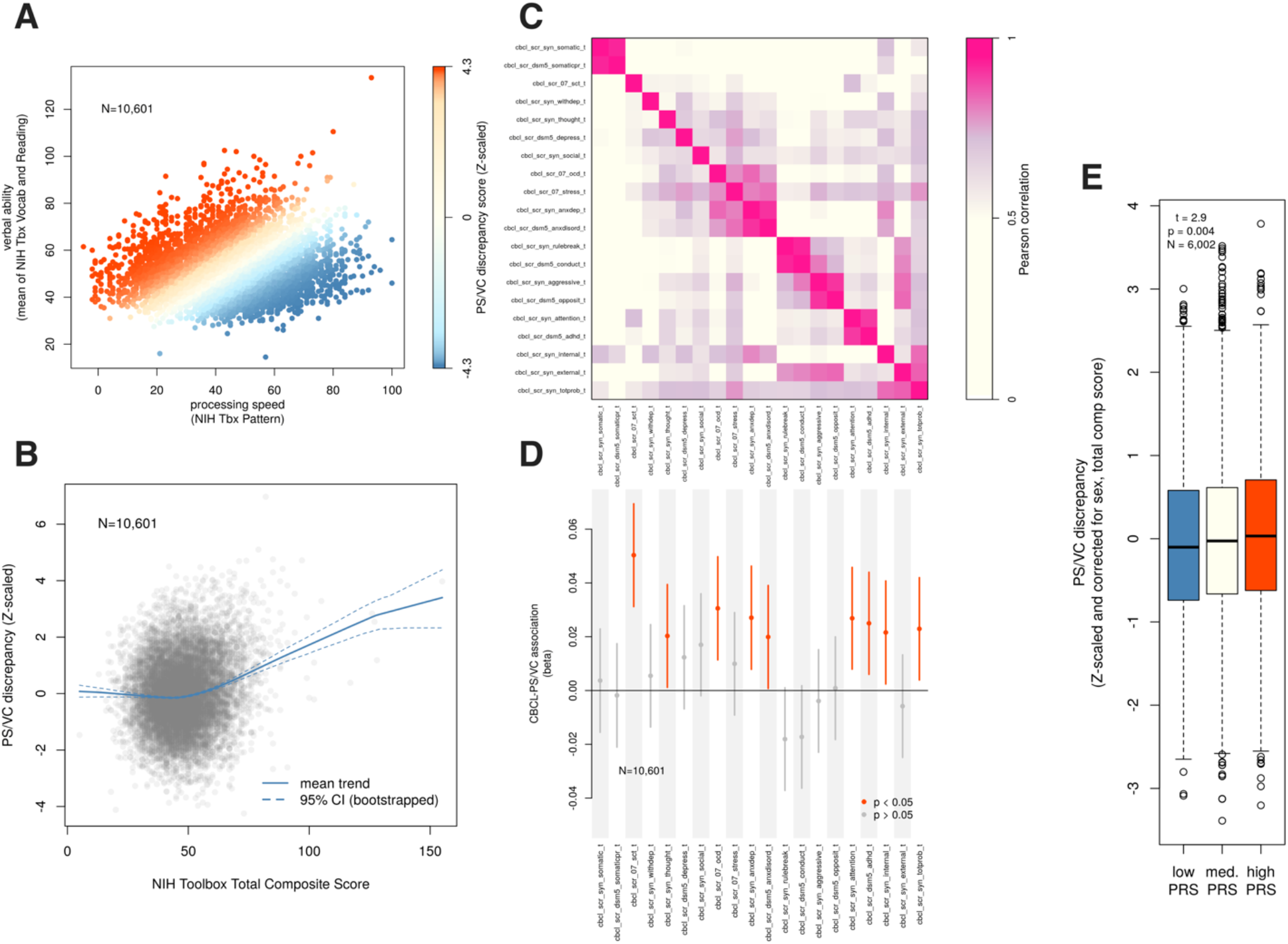
Generalization and polygenic association of the PS/VC discrepancy in the ABCD study. We used scores from the NIH Toolbox available from ABCD to represent verbal comprehension (mean of vocabulary and reading tasks) and processing speed (pattern task) (**A**). The difference between verbal and processing speed scores (PS/VC discrepancy) was Z-scaled and is depicted in color in panel A. The PS/VC discrepancy score is positively correlated with the NIH Toolbox total composite score, but this effect is driven largely by individuals at the higher end of this score (**B**), suggesting that the PS/VC discrepancy is particularly relevant for individuals with high overall cognitive ability. Computed subscales from the CBCL are correlated with each other to varying degrees, and our clustering identified loose super-clusters of internalizing and externalizing subscales (**C**). These CBCL subscales were tested for association with the PS/VC discrepancy score in order to test the generalization of our observations from the B-BC sample (**D**). We found evidence suggesting that PS/VC is significantly associated with anxiety and other internalizing symptoms, as well as attention problems, in agreement with our initial findings. Finally, we found that PS/VC is associated with increasing polygenic risk for autism (**panel E**, p = 0.004), in agreement with our initial observation of over-representation of autism in A5 (the PS-/VC+ cognitive archetype, see **Figure 3A**).

### Polygenic score analysis

Polygenic scores (PGS) for autism diagnostic status were computed in the ABCD sample (N=6,002) with LDPred2 [41] as described in [42]. Within ABCD, PGS were binned into three groups based on percentiles: depleted risk (20^th^ percentile and below), neutral risk (21^st^-79^th^ percentile), and high risk (80^th^ percentile and above). These three groups were coded with integer values 1, 2, and 3, respectively and were used as a quantitative explanatory variable. Sex and total composite score were used as additional covariates in a linear model predicting PS/VC discrepancy: PS_VC ~ sex + total_comp + PGS, where PS_VC is the PS/VC discrepancy score described above, total_comp is the NIH Toolbox total composite score, and PGS is the integer-coded polygenic score for autism. Tests of significance were performed using the fitted slope and its standard error (i.e., the summary() function in R).

## RESULTS

### A discrepancy between processing speed (PS) and verbal comprehension (VC) is associated with autism in the gifted clinical cohort

The central goal of our archetypal analysis was to uncover recurrent and distinct patterns of cognitive ability, and then determine whether any of those patterns are associated with autism diagnosis and other behaviors as observed from the BASC. This analysis is complementary to our diagnosis-driven analysis primarily because it utilizes an unsupervised learning approach and is agnostic to the diagnosis, which is only considered in a post-hoc analysis. Our analysis supported the presence of five cognitive archetypes in our sample (**Figure 3**). These archetypes correspond, approximately, to uniformly elevated ability (archetype 1, A1), generally lower ability with a specific strength in perceptual reasoning (archetype 2, A2), uniformly decreased ability (archetype 3, A3), strength in processing speed (PS) coupled with weakness in verbal comprehension (VC) (archetype 4, A4), and weakness in processing speed coupled with strength in verbal comprehension (archetype 5, A5). A total of 42 individuals did not meet criteria for exclusive assignment to any archetype and were considered “mixed”. Summary statistics for each of these archetypes are provided in Table 3. Notably, A4 and A5 are of particular interest because they are mirror images of each other (A4: PS+/VC− and A5: PS−/VC+). Further, these two cognitive archetypes show differential depletion and enrichment for autistic individuals, respectively (OR=0.39, 95% CI: [0.16,0.94] for A4 and OR=2.46, 95% CI: [1.5,4.1] for A5).

### Higher IQ delays autism diagnosis

In SPARK (N=17,634), individuals with parent-reported IQ of 120 or above were 5.2 times more likely than the low IQ group (IQ < 70) to be diagnosed after age six (OR=5.2 [4.7,5.2], p < 2.2×10^−16^), and twice as likely to be diagnosed in adulthood (i.e., age ≥ 18 years; OR=2.0 [1.2,3.5], p=0.01). See Figure 2. This parent-report data is limited by the lack of adults (most autistic adults who join SPARK do so without the involvement of their parents and also lack an option to self-report IQ in a way analogous to the parent-reported IQ), leading to over-convergence of diagnosis rates between the low and high groups in adulthood. To complement the analysis of the parent-reported IQ data, we also examined data from N=3,339 SPARK participants whose IQ scores were entered into their clinical records by medical providers in connection with a clinical IQ test, and subsequently gathered by SPARK recruitment centers who have consented access to these data. This analysis was in agreement with the overall trend observed in the parent-report data, but with much more pronounced effects: we found that individuals with IQ >=120 entered adulthood without a diagnosis at nearly 15 times the rate of their lower-IQ counterparts (OR=14.5 [2.8, 94.6], p=0.0005). Similarly, these high IQ individuals entered school age (i.e., age 6 years) without a diagnosis at more than nine times the rate of their lower-IQ peers (OR=9.2 [6.1,14.0], p < 2.2×10^−16^). Taken together, these converging results suggest that autistic individuals with exceptional ability spend proportionally less of their developing years without access to supports and services for their disabilities. Furthermore, the observation that an over-abundance of these high IQ autistic individuals do not receive a diagnosis until adulthood (and are therefore beyond the reach of autism experts in child psychiatry clinics) has ramifications for clinical practice in adult psychiatry, which often lacks specific expertise in neurodevelopmental conditions such as autism. This mismatch holds the potential for increased rates of misdiagnosis.

### Anxiety and attention problems increase with FSIQ specifically in autistic individuals in the gifted clinical cohort

The primary motivating question for our diagnosis-driven analysis was to understand whether increasing IQ might in some way be a liability for those with an autism diagnosis, when compared both with their non-autistic, high IQ peers, as well as their autistic but average IQ peers. This goal was operationalized by examining the interaction term in a linear model of functional outcomes (from the BASC) predicted by autism diagnosis, full-scale IQ, and their interaction (see Supplemental Table 2). Sex and age were included as additional covariates. As shown in Figure 3, we found that self-report of a sense of inadequacy had the strongest ASD*IQ interaction (beta=0.3, 95% CI:[0.15,0.45], p=7.1×10^−5^), indicating that autistic individuals show a 0.3 SD increase in this measure for each SD increase in FSIQ. Furthermore, anxiety was a point of confluence between self (beta=0.15, 95% CI: [0.01,0.29]), parent (beta=0.17, 95% CI: [0.05,0.29]), and teacher report (beta=0.14, 95% CI: [0.02,0.27]), though only parent report survived multiple testing correction at FDR < 0.05 (see Figure 3 A-D). Other significantly associated (all FDR<0.05) outcomes include parent reported functional communication (beta=-0.16, 95% CI: [-0.27,-0.05]) and attention problems (beta=0.16, 95% CI: [0.05,0.27]), teacher-reported school problems (beta=0.22, 95% CI: [0.07,0.37]), and self-reported hyperactivity (beta=0.22, 95% CI: [0.06,0.38]). To determine whether any of these associations would be considered potentially clinically meaningful, we dichotomized the BASC scores using the suggested threshold of >60 (or < 40 for reverse-scored variables). We found that teacher-reported school problems were nearly four times as likely to be clinically meaningful (i.e., by the thresholds indicated above) for an autistic individual with full-scale IQ of 120 vs. 100 (OR=4.2, 95% CI: [2.1,8.4]). Similarly, self-reported sense of inadequacy was three times as likely to be clinically relevant for autistic individuals at FSIQ 120 vs. 100 (OR=2.9, 95% CI: [1.4,5.9]), and parent reported functional communication issues more than twice as likely (OR=2.3, 95% CI: [1.6,3.5]).

### PS/VC discrepancy is predictive of internalizing, attention, and other problems in the gifted clinical cohort

Fourteen of 46 BASC measures were significantly associated with the PS/VC discrepancy at FDR < 0.05, and the full summary statistics of these tests are available in Supplemental Table 4. Briefly, parent and teacher reports of withdrawal, teacher-reported attention problems, parent- and teacher-reported atypicality, teacher-reported school problems, and teacher-reported depression were all significantly positively associated with PS/VC discrepancy (all FDR < 0.05, see Supplemental Table 4). Parent reports of activities of daily living, parent- and teacher-reported social skills, parent- and teacher-reported leadership skills, teacher-reported functional communication, and teacher-reported adaptability were all significantly negatively associated with PS/VC discrepancy all FDR < 0.05, see Supplemental Table 4). Together, these results paint a picture of the PS/VC discrepancy contributing to problems with attention, internalizing issues (withdrawal, depressive symptoms), poor leadership and social skills, and reduced adaptability, functional communication, and daily living skills. Notably absent are externalizing and somatization symptoms. Also noteworthy is the absence of association with any self-reported measures, which may suggest that those impacted by a PS/VC discrepancy may have little insight into the problems they face, while parents and teachers seem to be more attuned to these challenges.

### PS/VC discrepancy is predictive of internalizing and attention problems in both the gifted clinical cohort and the ABCD cohort

Nine of 20 available CBCL subscale scores showed a significant association at a nominal p < 0.05. Summary statistics for the PS/VC discrepancy score association are found in Supplemental Table 5 and the corresponding results are presented in **Figure 4**. The most striking association of the PS/VC discrepancy is with the “sluggish cognitive tempo” (SCT) subscale of the CBCL [40] (beta=0.05, t=5.2, p=2.6×10^−7^). This association, together with other significant associations with attention and ADHD-related subscales (attention: beta=0.03, t=2.8, p=0.005, ADHD: beta=0.03, t=2.6, p=0.009), suggest that PS/VC discrepancy is strongly associated with attention issues in non-clinical samples, in agreement with the findings from our analysis of the clinical sample. Further, we found that ABCD PS/VC is associated with a variety of internalizing symptoms, captured in the anxious/depressed, internalizing, and anxiety disorder subscales of the CBCL (all p < 0.05). Again, this connection of internalizing symptoms with PS/VC discrepancy is consistent with our findings in the clinical sample. Finally, a number of other subscale associations align with observations seen in the clinical sample: associations between PS/VC and OCD and thought problem subscales of the CBCL are congruent with the increased atypicality linked to A5 in the clinical sample, and the association with total problems (p < 0.05) may reflect the functional and daily living associations found in the clinical sample. Overall, these analyses suggest a general convergence in both clinical and non-clinical samples of the PS/VC cognitive discrepancy onto a combination of attention problems, internalizing symptoms, and atypicality.

### PS/VC discrepancy is associated with polygenic risk for autism in the ABCD cohort

Although the above analysis established the robustness of the behavioral associations with PS/VC discrepancy in an independent sample (ABCD), replicating the clinical sample’s observed enrichment (A5) and depletion (A4) of autism diagnosis is more challenging: ABCD does not have reliable diagnosis information that can be used to robustly identify participants with ASD. Instead, we appealed to the genetic data in ABCD to determine whether individuals with high polygenic risk for autism showed an elevated PS/VC. Indeed, autism PRS is significantly associated with PS/VC (beta=0.07, t=2.9, p=0.004), in agreement with our observation in the clinical sample of a significant excess of autism in A5 (and depletion in A4).

## DISCUSSION

High IQ is almost universally associated with better medical, behavioral, educational, social, and professional outcomes. These associations have influenced most clinicians’ and educators’ general expectations for their patients and students, and can lead to critical blind spots in situations where those expectations are violated. Consequently, there is a need to understand the specific contexts where high IQ can become a liability rather than an asset. The current study, which aims to address this need in the context of autism, included twice-exceptional students, i.e., autism coupled with very high IQ, as one of the subgroups, and expands upon previous research in several important ways. First, in addition to the twice-exceptional subgroup, there were three additional subgroups: average IQ + autism; gifted without autism, and average IQ without autism. Results from the current study corroborated previous findings[17–19] of exceptionally high verbal comprehension (VC) and exceptionally weak processing speed (PS). Furthermore, a larger and more diverse sample allowed for additional comparisons of main effects and interactions of IQ and psychosocial profiles from the four subsamples. In addition, the current investigation offered more robust support for the cognitive archetypes that are related to autism as well as for the impact of IQ on psychosocial profiles, especially for autistic individuals with high IQ. Finally, the use of ABCD data as an indicator of generalization of our findings extends the implications well beyond the clinical importance to include possible links to genetically meaningful biotypes.

In our more hypothesis-driven statistical analysis, we identified several key areas where increasing (full scale) IQ interacts with autism diagnosis to confer significant liabilities that diverge from the expectation based on the additive effects of diagnosis and IQ. Specifically, increasing IQ had a significant negative impact on sense of self-worth, anxiety, and attention on autistic individuals (Fig. 2). This surprising result is a stark departure from the main effect of IQ that ameliorates these symptoms and many others in the general population, and these observations are likely linked to increased rates of suicidality in 2e individuals [43]. These findings underscore the importance for educators, parents, and other caregivers to provide specific support in these areas, especially since they run counter to the general expectation for gifted individuals.

In our more hypothesis-driven statistical analysis, we identified several key areas where increasing (full scale) IQ interacts with autism diagnosis to confer significant liabilities that diverge from the expectation based on the additive effects of diagnosis and IQ. Specifically, increasing IQ had a significant negative impact on sense of self-worth, anxiety, and attention on autistic individuals (Fig. 2). This surprising result is a stark departure from the main effect of IQ that ameliorates these symptoms and many others in the general population, and these observations are likely linked to increased rates of suicidality in 2e individuals [43]. These findings underscore the importance for clinicians, educators, parents, and other caregivers to provide specific support in these areas, especially since they run counter to the general expectation for gifted individuals.

Our unsupervised learning analysis discovered several cognitive archetypes (i.e., recurrent patterns of strength and weakness across IQ index scores) that showed significant associations, both positive and negative, with autism diagnosis (Fig. 3). Although autism is defined by DSM-V criteria based on behavior, these findings suggest that basic cognitive patterns, which are more amenable to measurement than behavior, may be indicative of the neural processes and mechanisms underlying autism. Specifically, two archetypes, A4 and A5, were mirror images of each other: A4 showing a relative strength in PS and a deficit in VC, and A5 showing the opposite, a deficit in PS and a strength in VC. A4 was significantly depleted for autistic individuals, while A5 showed an enrichment of comparable effect size. These two archetypes suggest an axis defined by changes in PS relative to VC, and vice versa, that also indexes autism risk. As an additional analysis to test the hypothesis of a link between autism risk and relative differences in PS and VC, we utilized the ABCD study, a general population sample that includes genetic, cognitive, and behavioral data (Fig. 4). We found a modest but significant relationship, in a direction in agreement with our clinical sample findings, between a polygenic score for autism and a PS/VC differential (derived from NIH Toolbox scores). Furthermore, the PS/VC differential predicted behavioral phenotypes in ABCD, relating to attention and internalizing symptoms, that are similar to those observed in the clinical sample. Taken together, these results from multiple cohorts point toward a “cognitive divergence” between PS and VC that also indexes autism risk in both clinical and general population samples. This constellation of findings points to a mechanism that underlies at least part of the autism spectrum, and may be a key in defining phenotypes that are more likely to exhibit specific patterns of genetic risk of the kind that have so far eluded attempts to disentangle autism from more general neurodevelopmental disabilities[44]. Importantly, while previous studies have found that deficits in PS are associated with a variety of diagnoses [45–47], including autism [48–50], our observation of the “mirror” phenotype, i.e., high PS and low VC, apparently having a protective effect against autism, is novel. Coupling these two extremes together into a PS/VC tradeoff axis may yield critical new insights into the cognitive foundations of autism.

The results of this study speak to the real and often unappreciated challenges faced by autistic individuals with extremely high ability. A recent commission report on autism [51] advocated for the use of a new term, “profound autism”, to describe individuals who require substantial daily living support and who need near-constant care and supervision that will continue into adulthood. The commission lamented that these individuals and their families are “at risk of being marginalized by a focus on more able individuals”. We concur that the risk of marginalization is a critical issue, and recognize that more can and should be done to help these individuals and families who have significant needs. However, our data offer important additional insight that seems to contrast with the commission’s assertion: we showed that twice-exceptional (“more able”) individuals are marginalized by delayed diagnosis and corresponding lack of support, compared to individuals with average or low IQ. In other words, autistic individuals with high IQ are at risk of having their disability and corresponding mental health concerns minimized rather than recognized. The needs and disabilities of these groups may differ dramatically in their nature, but codifying language that implies a basic difference in intensity or priority (i.e., “profound”) runs the risk of perpetuating the minimization of disabilities that are bound together with extreme ability.

Although our findings point to important conclusions about the relationship between high IQ and autism, there are several key limitations that should be taken into account when interpreting these results. First, our study relies on symptoms reported by affected individuals, their parents, and educators. While these are highly relevant perspectives, it remains unclear whether twice-exceptional individuals suffer more clinically significant complications, e.g., increased rates for hospitalization or suicidality. A better understanding of these high-impact outcomes is the focus of ongoing research. On the other hand, our primary cohort is a clinical sample, and might not completely reflect 2e individuals who were not clinically assessed. Our analysis of the ABCD study does however suggest generalizability between a PS/VC divergence and autism-like symptoms. Furthermore, the lack of either genetic or neuroimaging data in our clinical sample prevents us from developing deeper mechanistic insight into the cognitive and behavioral phenomena we observed. Until such data are collected, we are limited to making these connections in larger, general population samples (like ABCD), which do not have an enrichment for either gifted or 2e individuals. Finally, twice-exceptionality does not imply or require an autism diagnosis specifically, and it is unclear how twice-exceptionality that involves other diagnoses beyond autism, such as ADHD or specific learning disorders, compare and contrast with the twice-exceptionality we describe here.

The results we present here argue a strong case for further research on the causes and consequences of disability linked to high IQ in autistic and other neurodiverse individuals. These twice-exceptional individuals suffer profoundly from crippling anxiety, attention problems, and depressive symptoms that put them at substantially increased risk for suicide. The more focus researchers and clinicians devote to these issues, especially on earlier diagnosis and support, the more society will benefit from the unlocked potential of these gifted yet disabled individuals.

## Data Availability

Data will be made publicly available upon acceptance of manuscript to a peer-reviewed journal.

https://nda.nih.gov/abcd

## ACKNOWLEDGEMENTS

We are grateful to all of the individuals and families participating in SPARK, the SPARK clinical sites, and SPARK staff. We appreciate obtaining access to genetic and phenotypic data for SPARK data on SFARI Base. We are also appreciative of the individuals and families in the ABCD study and the clinical sample.

This work was supported by a grant from the Simons Foundation (SFARI Explorer 594788 to JJM). This work was also supported by the University of Iowa Hawkeye Intellectual and Developmental Disabilities Research Center P50 HD103556 (TA and Lane Strathearn, multi-PIs). Additional funding for this work came from the Provost’s Office and Department of Psychiatry at the University of Iowa, the Roy J. Carver Charitable Trust, the University of Iowa Institute for Clinical and Translational Science (UL1TR002537), the National Institutes of Health through a Predoctoral Training Grant (T32GM008629 to LC and TT), and Research Grant DC014489 to JJM.

## References

1. Andreasen, N.C., Creativity and mental illness: prevalence rates in writers and their first-degree relatives. Am J Psychiatry, 1987. 144(10): p. 1288–92.

2. Maier, T., [Psychosis, language and literature]. Nervenarzt, 1999. 70(5): p. 438–43.

3. Thorup, A.A.E., et al., Exploring protective and risk factors in the home environment in high-risk families - results from the Danish High Risk and Resilience Study-VIA 7. BMC Psychiatry, 2022. 22(1): p. 100.

4. Smeland, O.B., et al., Genome-wide analysis reveals extensive genetic overlap between schizophrenia, bipolar disorder, and intelligence. Mol Psychiatry, 2020. 25(4): p. 844–853.

5. Zabaneh, D., et al., A genome-wide association study for extremely high intelligence. Mol Psychiatry, 2018. 23(5): p. 1226–1232.

6. Savage, J.E., et al., Genome-wide association meta-analysis in 269,867 individuals identifies new genetic and functional links to intelligence. Nat Genet, 2018. 50(7): p. 912–919.

7. Grove, J., et al., Identification of common genetic risk variants for autism spectrum disorder. Nat Genet, 2019. 51(3): p. 431–444.

8. Clarke, T.K., et al., Common polygenic risk for autism spectrum disorder (ASD) is associated with cognitive ability in the general population. Mol Psychiatry, 2016. 21(3): p. 419–25.

9. Verhoef, E., et al., Discordant associations of educational attainment with ASD and ADHD implicate a polygenic form of pleiotropy. Nat Commun, 2021. 12(1): p. 6534.

10. Radua, J., et al., What causes psychosis? An umbrella review of risk and protective factors. World Psychiatry, 2018. 17(1): p. 49–66.

11. Eling, P., History of Neuropsychological Assessment. Front Neurol Neurosci, 2019. 44: p. 164–178.

12. Gale, C.R., et al., Intelligence in early adulthood and subsequent hospitalization for mental disorders. Epidemiology, 2010. 21(1): p. 70–7.

13. Williams, D.L., G. Goldstein, and N.J. Minshew, Neuropsychologic functioning in children with autism: further evidence for disordered complex information-processing. Child Neuropsychol, 2006. 12(4-5): p. 279–98.

14. Gomez, R., A. Vance, and S.D. Watson, Structure of the Wechsler Intelligence Scale for Children - Fourth Edition in a Group of Children with ADHD. Front Psychol, 2016. 7: p. 737.

15. Mayes, S.D. and S.L. Calhoun, WISC-IV and WIAT-II profiles in children with high-functioning autism. J Autism Dev Disord, 2008. 38(3): p. 428–39.

16. Gale, C.R., et al., Is bipolar disorder more common in highly intelligent people? A cohort study of a million men. Mol Psychiatry, 2013. 18(2): p. 190–4.

17. Foley-Nicpon, M. and S. Lee, Disability research in counseling psychology journals: a 20-year content analysis. J Couns Psychol, 2012. 59(3): p. 392–8.

18. Assouline, S.G., M. Foley Nicpon, and L. Dockery, Predicting the academic achievement of gifted students with autism spectrum disorder. J Autism Dev Disord, 2012. 42(9): p. 1781–9.

19. Doobay, A.F., et al., Cognitive, Adaptive, and Psychosocial Differences Between High Ability Youth With and Without Autism Spectrum Disorder. Journal of Autism and Developmental Disorders, 2014. 44(8): p. 2026–2040.

20. Beckmann, E. and A. Minnaert, Non-cognitive Characteristics of Gifted Students With Learning Disabilities: An In-depth Systematic Review. Front Psychol, 2018. 9: p. 504.

21. Reis, S.M., S.M. Baum, and E. Burke, An Operational Definition of Twice-Exceptional Learners: Implications and Applications. Gifted Child Quarterly, 2014. 58(3): p. 217–230.

22. Devlin, B., M. Daniels, and K. Roeder, The heritability of IQ. Nature, 1997. 388(6641): p. 468–71.

23. Bai, D., et al., Association of Genetic and Environmental Factors With Autism in a 5-Country Cohort. JAMA Psychiatry, 2019. 76(10): p. 1035–1043.

24. Lee, J.J., et al., Gene discovery and polygenic prediction from a genome-wide association study of educational attainment in 1.1 million individuals. Nat Genet, 2018. 50(8): p. 1112–1121.

25. Feliciano, P., et al., SPARK: A US Cohort of 50,000 Families to Accelerate Autism Research. Neuron, 2018. 97(3): p. 488–493.

26. Lisdahl, K.M., et al., Adolescent brain cognitive development (ABCD) study: Overview of substance use assessment methods. Dev Cogn Neurosci, 2018. 32: p. 80–96.

27. Marees, A.T., et al., A tutorial on conducting genome-wide association studies: Quality control and statistical analysis. Int J Methods Psychiatr Res, 2018. 27(2): p. e1608.

28. Purcell, S., et al., PLINK: a tool set for whole-genome association and population-based linkage analyses. Am J Hum Genet, 2007. 81(3): p. 559–75.

29. R Core Team, R: A Language and Environment for Statistical Computing. 2013, Vienna, Austria: R Foundation for Statistical Computing.

30. Lemieux Perreault, L.P., et al., genipe: an automated genome-wide imputation pipeline with automatic reporting and statistical tools. Bioinformatics, 2016. 32(23): p. 3661–3663.

31. Delaneau, O., J. Marchini, and J.F. Zagury, A linear complexity phasing method for thousands of genomes. Nat Methods, 2011. 9(2): p. 179–81.

32. Howie, B.N., P. Donnelly, and J. Marchini, A flexible and accurate genotype imputation method for the next generation of genome-wide association studies. PLoS Genet, 2009. 5(6): p. e1000529.

33. Sattler, J.M., Assessment of Children: Cognitive Foundations and Applications. 2018, La Mesa, CA: Jerome Sattler Publications.

34. Wechsler, D., Wechsler Intelligence Scale for Children (5th Edition). 2014, San Antonio, TX: NCS Pearson.

35. Wechsler, D., Wechsler Intelligence Scale for Children (5th Edition) Technical and Interpretive Manual. 2014, San Antonio, TX: NCS Pearson.

36. Reynolds, C.R. and R.W. Kamphaus, BASC 3: Behavior Assessment System for Children (3rd Edition) Manual. 2015, Bloomington, MN: Psych-Corp, NCS Pearson.

37. Benjamini, Y. and Y. Hochberg, Controlling the False Discovery Rate - a Practical and Powerful Approach to Multiple Testing. Journal of the Royal Statistical Society Series B-Methodological, 1995. 57(1): p. 289–300.

38. Cutler, A. and L. Breiman, Archetypal Analysis. Technometrics, 1994. 36(4): p. 338–347.

39. Casey, B.J., et al., The Adolescent Brain Cognitive Development (ABCD) study: Imaging acquisition across 21 sites. Dev Cogn Neurosci, 2018. 32: p. 43–54.

40. Achenbach, T.M., et al., Multicultural assessment of child and adolescent psychopathology with ASEBA and SDQ instruments: research findings, applications, and future directions. Journal of Child Psychology and Psychiatry, 2008. 49(3): p. 251–275.

41. Prive, F., J. Arbel, and B.J. Vilhjalmsson, LDpred2: better, faster, stronger. Bioinformatics, 2020.

42. Thomas, T.R., et al., Clinical autism subscales have common genetic liability that is heritable, pleiotropic, and generalizable to the general population. medRxiv, 2022: p. 2021.08.30.21262845.

43. Casten, L.G., et al., The combination of autism and exceptional cognitive ability increases risk for suicidal ideation. medRxiv, 2022: p. 2022.02.17.22271086.

44. Myers, S.M., et al., Insufficient Evidence for “Autism-Specific” Genes. Am J Hum Genet, 2020. 106(5): p. 587–595.

45. Braaten, E.B., et al., Characteristics of child psychiatric outpatients with slow processing speed and potential mechanisms of academic impact. Eur Child Adolesc Psychiatry, 2020. 29(10): p. 1453–1464.

46. Kramer, E., et al., Diagnostic Associations of Processing Speed in a Transdiagnostic, Pediatric Sample. Sci Rep, 2020. 10(1): p. 10114.

47. Eack, S.M., et al., Commonalities in social and non-social cognitive impairments in adults with autism spectrum disorder and schizophrenia. Schizophr Res, 2013. 148(1-3): p. 24–8.

48. Travers, B.G., et al., Longitudinal processing speed impairments in males with autism and the effects of white matter microstructure. Neuropsychologia, 2014. 53: p. 137–45.

49. Haigh, S.M., et al., Processing Speed is Impaired in Adults with Autism Spectrum Disorder, and Relates to Social Communication Abilities. Journal of Autism and Developmental Disorders, 2018. 48(8): p. 2653–2662.

50. Oliveras-Rentas, R.E., et al., WISC-IV Profile in High-Functioning Autism Spectrum Disorders: Impaired Processing Speed is Associated with Increased Autism Communication Symptoms and Decreased Adaptive Communication Abilities. Journal of Autism and Developmental Disorders, 2012. 42(5): p. 655–664.

51. Lord, C., et al., The Lancet Commission on the future of care and clinical research in autism. Lancet, 2022. 399(10321): p. 271–334.

